# The impact of a conditional financial incentive on linkage to HIV care: Findings from the HITS cluster randomized clinical trial in rural South Africa

**DOI:** 10.1101/2024.03.15.24304278

**Authors:** Hae-Young Kim, Maxime Inghels, Thulile Mathenjwa, Maryam Shahmanesh, Janet Seeley, Phillippa Matthews, Nuala McGrath, Oluwafemi Adeagbo, Dickman Gareta, H. Manisha Yapa, Thembelihle Zuma, Adrian Dobra, Till Bärnighausen, Frank Tanser

**Affiliations:** New York University School of Medicine, New York, USA; Africa Health Research Institute, KwaZulu-Natal, South Africa; Lincoln International Institute for Rural Health, University of Lincoln, Lincoln, United Kingdom; Centre Population et Développement (UMR 196 Paris Descartes – IRD), SageSud (ERL INSERM 1244), Institut de Recherche pour le Développement, Paris, France; Institute for Global Health, University College London, London, United Kingdom; London School of Hygiene and Tropical Medicine, London, United Kingdom; University of Glasgow, Glasgow, United Kingdom; University of Southampton, Southampton, United Kingdom; University of Johannesburg, Johannesburg, South Africa; College of Public Health, University of Iowa, USA; The Kirby Institute, University of New South Wales Sydney, Sydney, Australia; Sydney Infectious Diseases Institute, University of Sydney, Sydney, Australia; University of Washington, Washington, USA; University College London Interaction Centre, University College London, London, United Kingdom; Heidelberg Institute of Global Health, University of Heidelberg, Heidelberg, Germany; Centre for Epidemic Response and Innovation, School for Data Science and Computational Thinking, Stellenbosch University, Stellenbosch, South Africa; South African DSI-NRF Centre of Excellence in Epidemiological Modelling and Analysis (SACEMA), Stellenbosch University, Stellenbosch, South Africa; School of Nursing and Public Health, University of KwaZulu-Natal, Durban, South Africa

**Keywords:** <u>Keywords</u>: Linkage to HIV care, financial incentives, randomized controlled trial, South Africa

## Abstract

**Introduction:** HIV elimination requires innovative approaches to ensure testing and immediate treatment provision. We investigated the effectiveness of conditional financial incentives on increasing linkage to HIV care in a 2×2 factorial cluster randomized controlled trial-Home-Based Intervention to Test and Start (HITS) - in rural South Africa.

**Methods:** Of 45 communities in uMkhanyakude, KwaZulu-Natal, 16 communities were randomly assigned to the arms to receive financial incentives for home-based HIV counseling and testing (HBHCT) and linkage to care within 6 weeks (R50 [US$3] food voucher each) and 29 communities to the arms without financial incentives. We examined linkage to care (i.e., initiation or resumption of antiretroviral therapy after >3 months of care interruption) at local clinics within 6 weeks of a home visit, the eligibility period to receive the second financial incentive. Linkage to care was ascertained from individual clinical records. Intention-to-treat analysis (ITT) was performed using modified Poisson regression with adjustment for receiving another intervention (i.e., male-targeted HIV-specific decision support app) and clustering of standard errors at the community level.

**Results:** Among 13,894 eligible men (i.e., ≥15 years and resident in the 45 communities), 20.7% received HBHCT, which resulted in 122 HIV-positive tests. Of these, 27 linked to care within 6 weeks of HBHCT. Additionally, of eligible men who did not receive HBHCT, 66 linked to care. In the ITT analysis, the proportion of linkage to care among men did not differ in the arms which received financial incentives and those without financial incentives (adjusted Risk Ratio [aRR]=0.78, 95% CI: 0.51-1.21). Among 19,884 eligible women, 29.1% received HBHCT, which resulted in 375 HIV-positive tests. Of these, 75 linked to care. Among eligible women who did not receive HBHCT, 121 linked to care within 6 weeks. Women in the financial incentive arms had a significantly higher probability of linkage to care, compared to those in the arms without financial incentives (aRR=1.50; 95% CI: 1.03-2.21).

**Conclusion:** While a small once-off financial incentive did not increase linkage to care among men during the eligibility period of 6 weeks, it significantly improved linkage to care among women over the same period.

Clinical Trial Number: ClinicalTrials.gov # NCT03757104

## INTRODUCTION

Antiretroviral therapy (ART) has dramatically reduced HIV-related morbidity and mortality as well as the risk of HIV transmission among people living with HIV (PLWH).^1–7^ To maximize the benefits of ART, it is critical to diagnose PLWH early, link and retain them in care. However, a substantial proportion of PLWH remains undiagnosed or not linked to care. In South Africa, which is home to over 7 million PLWH and 210,000 new infections in 2021, 94% of PLWH knew their HIV status, but only 74% of them linked to care and received ART.^8^

Home-based HIV counseling and testing (HBHCT) is an effective way to reach the undiagnosed or hard-to-reach population in rural^9–13^ and urban^12^ settings in sub-Saharan Africa (SSA).^14^ HBHCT is highly acceptable and has increased the uptake of HIV testing in SSA.^14^ However, several studies have shown that linkage to care following HBHCT remains suboptimal,^15^ particularly among newly diagnosed individuals.^10,16^ For example, in HPTN 071 (PopART), a community-randomized trial of a combination HIV prevention package, only 36% and 66% of newly diagnosed individuals initiated ART by 3 and 12 months, respectively.^17^ Especially, men are more likely to delay their linkage to care after diagnosis,^17,18^ due to barriers such as perceptions about the treatment, social stigma or gender norms.^16,19^

Various interventions have been implemented to improve linkage to care in SSA, including the provision of financial incentives.^20,21^ Conditional financial incentives had shown mixed results in increasing linkage to care. Several studies conducted in SSA have reported that financial incentives, when combined with intervention strategies such as point-of-care testing, did not significantly improve linkage to care.^22–30^ However, to our knowledge, no study has examined the impact of multi-stage financial incentives and their impact on linkage to care following HBHCT in SSA.

We conducted a 2×2 factorial cluster randomized controlled trial-Home-Based Intervention to Test and Start (HITS) - in 45 rural communities in uMkhanyakude district of KwaZulu Natal, South Africa.

We previously reported findings on the impact of conditional financial incentives and the app-based system for informed decision-making, EPIC-HIV, on the uptake of HBHCT.^31,32^ Here, we report the impact of financial incentive on linkage to care among men and women.

## METHODS

### Setting

The trial was nested within AHRI’s Health and Demographic Surveillance System (HDSS) and facilitated linkage to care in uMkhanyakude district of Northern KwaZulu Natal.^33^ The estimated HIV prevalence in the study area was estimated as 19% in men and 40% in women in 2018.^1^ Since 2003, AHRI has conducted population-based HIV surveillance within AHRI’s HDSS. The HIV surveillance is annually conducted among all residents aged ≥15 years to collect data on sexual behavior and general health as well as dried blood spots (DBS) samples for anonymized HIV testing after obtaining informed consent.^34,35^ Since 2017, rapid HIV testing with immediate results has been offered as part of the HIV surveillance during household visits.

### Trial Design

Between February and December 2018, we implemented a 2×2 factorial cluster-randomized trial among a population of 37,028 residents aged ≥15 years across 45 clusters within AHRI’s ongoing population-based annual HIV testing platform (Figure 1).^36^ The trial offered two interventions: financial incentives and a male-targeted app-based system for informed decision-making, called EPIC-HIV (Empowering People through Informed Choices for HIV). Over the entire study duration, 8 communities received only financial incentives, 8 communities received only EPIC-HIV, 8 communities received both interventions, and 21 communities received standard of care. While both males and females were eligible to receive financial incentives, only males were eligible to receive EPIC-HIV. Randomization was conducted to ensure balance across the arms using stratified sampling at the community level based on the HIV incidence among young females aged 15-30 years. The implementation and acceptance of the HITS intervention were evaluated using a process evaluation, utilizing post-intervention satisfaction surveys^37^ as well as focus group discussion and in-depth interviews among study participants, fieldworkers, and health professionals. Results of the process evaluation have been published elsewhere.^38^ The trial was registered at the National Institute of Health (ClinicalTrials.gov # NCT03757104), and the complete trial protocol was published elsewhere.^36^

**Figure 1.**
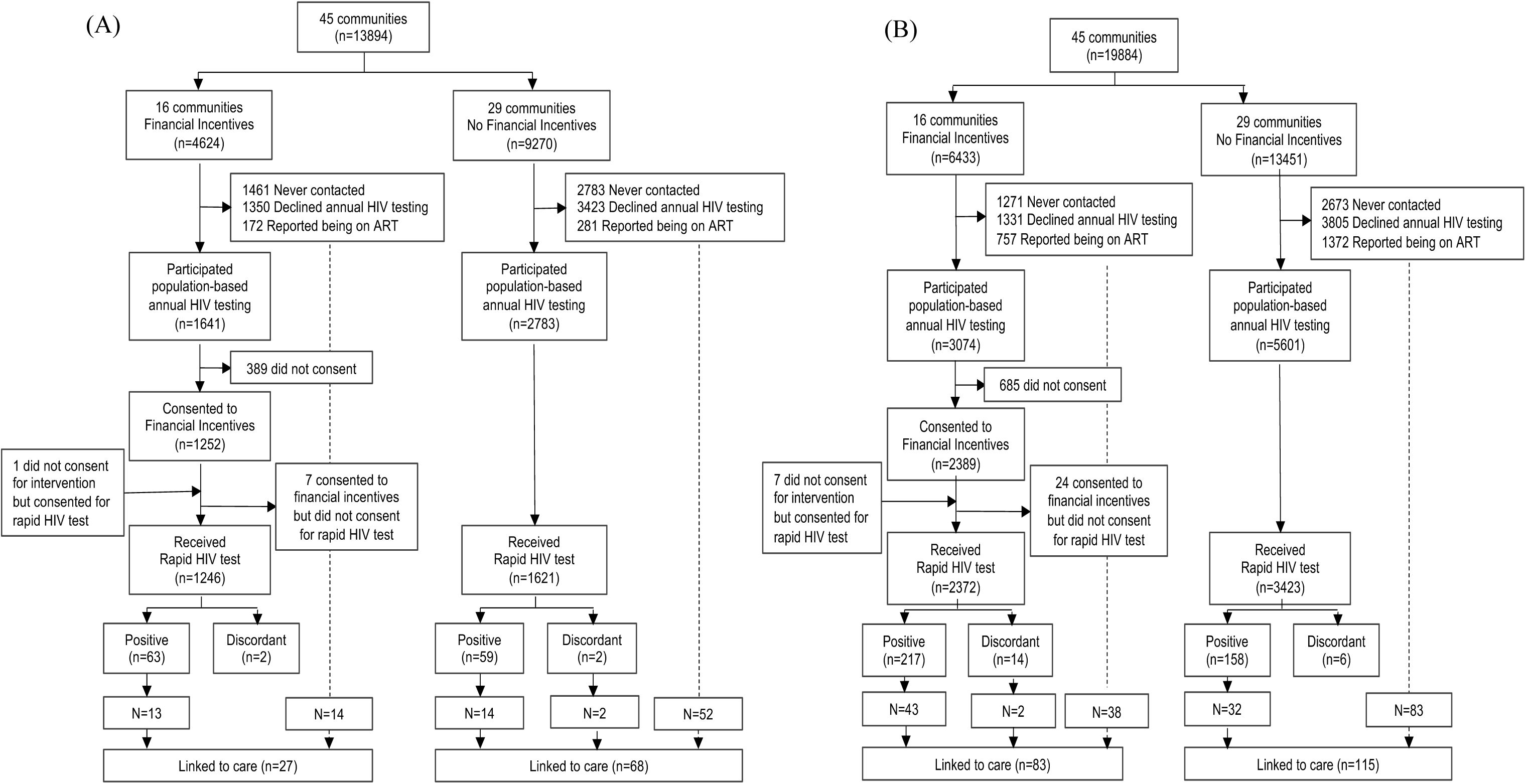
Flow diagram for the HITS cluster-randomized controlled trial and linkage to care within the voucher eligibility period of 6 weeks of a home visit among (A) men and (B) women. Flow diagram shows individual flow through each stage of the HITS trial in the arms with and without financial incentives. The dashed line indicates linkage to care within 6 weeks among those who were never contacted, declined annual HIV testing, or did not consent for interventions.

**Figure 2.**
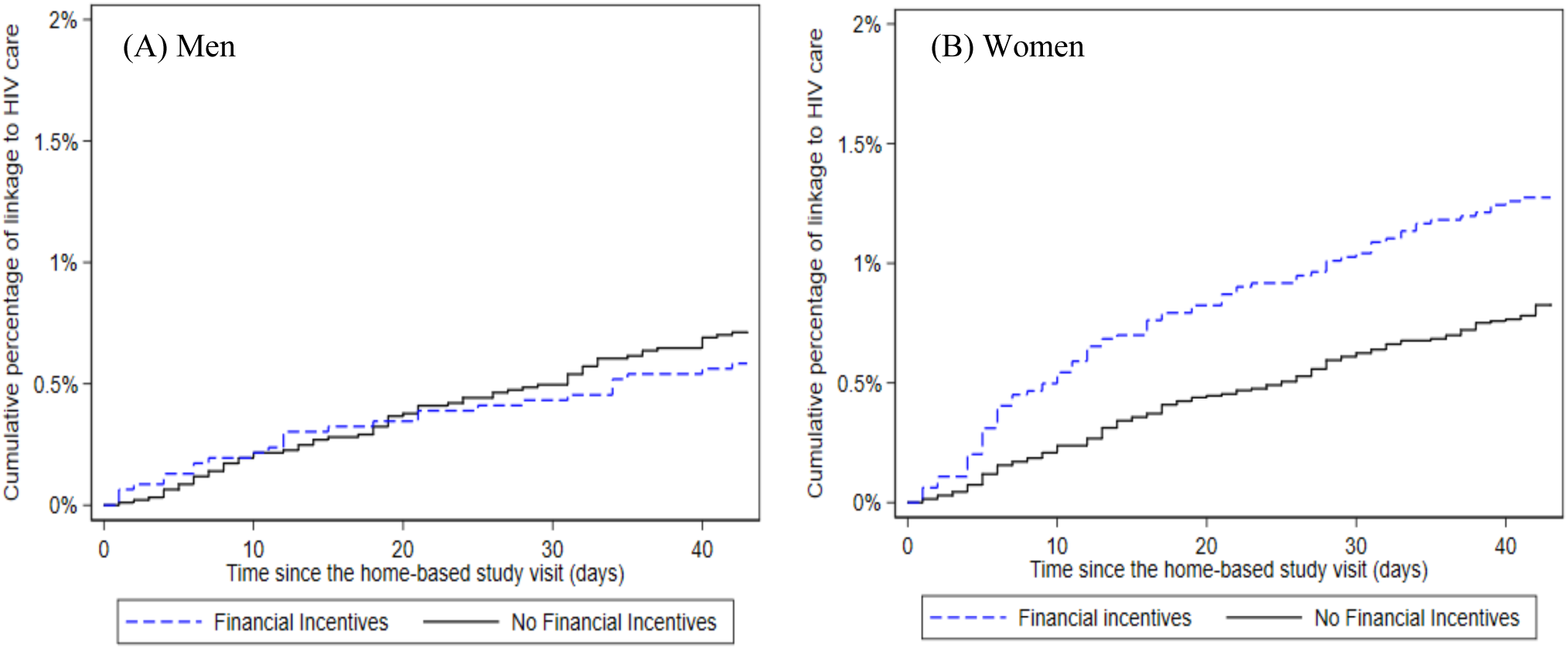
Kaplan Meir curves for linkage to HIV care by the voucher eligibility period of 6 weeks after the home-based study visit among (A) men and (B) women. The solid black lines represent the arms without financial incentives, and the dashed blue lines the financial incentive arms.

### Standard of care for HBHCT and linkage to care

As part of the AHRI population-based annual HIV surveillance, field workers who are trained in HIV counselling and testing conduct rapid point-of-care HIV tests with immediate results in accordance with the South African national guidelines. When individuals are not available during the initial contact attempt to complete the HIV surveillance, they are revisited twice more during normal working hours within the same week. Subsequently, a tracking team attempts to contact participants and make three additional home visits during evenings or weekends before considering them as non-contact. Participants found to be HIV-positive are encouraged to link to care within 7-10 days of the HIV test date and are offered a referral slip for an appointment to receive HIV care at one of the 10 local clinics. In these clinics, AHRI has set up a system to record clinic visits and reasons for attendance for all consenting individuals presenting at the clinics. Individuals who consent for facilitated linkage and have not linked to care within two weeks of the HIV test date receive a single Short Message Service (SMS) message as a reminder. If these individuals do not link to care within an additional two weeks, a trained nurse contacts them by telephone to discuss any concerns and encourages them to link to care.

### Interventions

The interventions were delivered in a two-stage scheme for HIV testing and linkage to care. In the arms receiving financial incentives, participants were offered a R50 (US ∼$3) food voucher for a local supermarket conditional on their participation in rapid HIV testing. Second, participants who tested HIV-positive were offered another R50 food voucher if they visited any of the 10 primary health clinics in the AHRI HIV surveillance area to seek HIV treatment within 6 weeks of the positive HIV test date. The male-targeted HIV-specific decision support application was implemented and offered via a tablet and offered in two versions (EPIC-HIV 1 and EPIC-HIV 2). EPIC-HIV 1 was provided to men prior to the offer of HIV testing. It aimed to support their decision-making regarding whether to take a rapid HIV test or not and to facilitate linkage to care if tested HIV-positive. Participants who did not link to HIV care within a month of a positive HIV test were revisited by a study tracker and offered the second application (EPIC-HIV 2), which is designed to address barriers to seeking HIV treatment and encourage them to link to HIV care. The application development is described in detail elsewhere.^36,39^ Individuals who did not receive home-based HIV testing, including those who were not contacted or declined annual HIV testing, were not eligible to receive the financial incentive upon linkage to care.

### Outcomes

In this study, we report the proportion of individuals diagnosed with HIV following HBHCT and the probability of linkage to care within 6 weeks after HBHCT, the eligibility period to receive the second financial incentive. The conditional financial incentive for linkage to care was only provided if a person tested HIV-positive at HBHCT linked to care within 6 weeks after HBHCT. Linkage to care was defined as ART initiation or resumption and captured using two sources of data collection, covering all study participants including those who were never contacted or declined annual HIV testing during the study visit. First, clinical research assistants stationed at 10 local clinics within the surveillance area and recorded clinical information for all visiting patients, including clinic visit dates, reasons for clinic attendance, and participation in the AHRI HDSS. This information was captured into an electronic data collection system called AHRILink, where the database for each of the 10 clinics was linked to a central database through an ongoing replication process. Second, through a memorandum of agreement with the South African Department of Health, the AHRI population-based program is linked with the clinical records of patients registered in the local public HIV Treatment and Care Programme at the Hlabisa district hospital and 17 primary health care clinics within the Hlabisa health sub-district using the TIER.Net. TIER.Net is a three-tiered electronic patient management system used in public clinics in South Africa for monitoring and evaluating HIV care and treatment for all patients receiving ART. The TIER.Net system was implemented in uMkhanyakude district in 2013. Patient records from all visits prior to 2013 were back-captured into the system from AHRI’s previous HIV care clinical database between 2004 and 2012.^40^ The TIER.Net database is linked with the AHRI surveillance database based on personal identifiers using algorithms developed at AHRI. All patient clinic visits for ART initiation and follow-up after the home-based study visits were confirmed through Tier.Net and AHRILink. ART initiation was defined as being newly prescribed ART without any prior record of ART initiation. ART resumption was defined as re-initiating ART after >90 days of care interruption as ascertained through AHRILink and/or TIER.Net.

### Statistical methods

The primary analysis was conducted using the intent-to-treat (ITT) analysis for all men and women randomized at the community level. We examined the outcome as a binary variable using modified Poisson regression, adjusting for clustering of standard errors at the community level. All outcomes among males were also adjusted for the provision of the EPIC intervention. We estimated the time to linkage to care within 6 weeks for each intervention arm using Kaplan-Meir survival curves. The log rank test was used to compare the differences in linkage to care in the arms with and without financial incentives. All analyses were conducted among men and women, separately, using STATA 15.1 (StataCorp) and R 4.0.3.

### Ethics statement

The study protocols for the AHRI’s population-based HIV testing platform and HITS intervention were approved by the Biomedical Research Ethics Committee of the University of KwaZulu-Natal (BE290/16 and BFC398/16).^36^ Permission for the trial was obtained from the KwaZulu-Natal Department of Health, South Africa. Participation in the HIV surveillance and HITS trial is completely voluntary. Individuals may choose not to answer and/or participate in any component of the HIV surveillance and to withdraw at any time. Written informed consent was sought from individuals aged ≥ 18 years, and parental or guardian consent with child assent for individuals of 15-17 years old were obtained.

## RESULTS

### Participants and recruitment flow

All 15,485 men living in the 45 clusters in the study area were initially considered eligible for the trial. Study participants were enrolled between February 2018 and December 2018. Of these, 1,591 died or out-migrated. Of the remaining 13,894 eligible men, 4,244 (30.6%) could not be contacted (mainly due to absence at the time of HIV testing despite several attempts to follow up), 4,773 (34.4%) declined to participate in the annual population-based HIV testing in 2018, and 453 (3.3%) self-reported being on ART, thus resulting in 4,424 (31.8% of the resident population) who participated in the population-based HIV testing (Figure 1). The flow diagram through each stage of the HITS trial by the 2×2 intervention arms among men is available in Figure S1. Randomization successfully achieved balance regarding HIV prevalence and sociodemographic variables in the arms with vs. without financial incentives, except for the area of residency (Table 1).

**Table 1.**
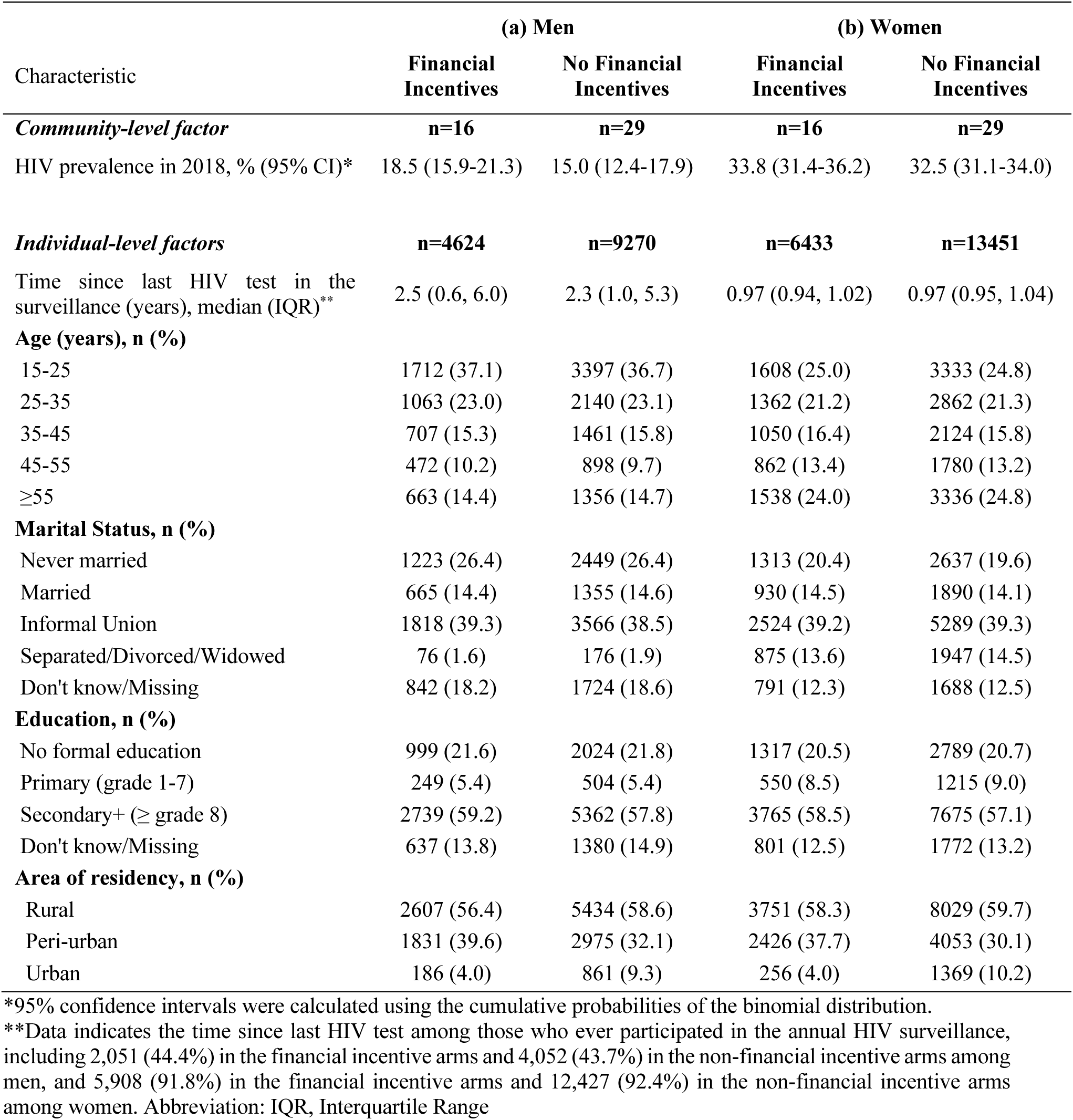
Baseline characteristics of communities and individuals among men and women in the arms with and without financial incentives.

**Table 2.** Linkage to HIV care within the voucher eligibility period of 6 weeks among men and women in the arms with and without the financial incentives.

|  | N | Linked to HIV care<br>within 6 weeks, n(%) | Risk Ratio* (95% CI) | p-value |
| --- | --- | --- | --- | --- |
| **Men** |  |  |  |  |
| Financial incentives | 4624 | 27 (0.58) | 0.78 (0.50-1.21) | 0.27 |
| No financial incentives | 6746 | 68 (0.73) | Ref |  |
| **Women** |  |  |  |  |
| Financial incentives | 6433 | 83 (1.29) | 1.50 (1.03-2.21) | 0.04 |
| No financial incentives | 13451 | 115 (0.85) | Ref |  |

\*Risk ratio accounts for clustering within communities using a random-intercept log-Poisson regression with empirical standard error estimates. It was also adjusted for the provision of EPIC-HIV provided as part of the 2x2 cluster randomized trial design.

Of 21,545 women residing in the 45 clusters within the study area initially considered eligible for inclusion, 1,661 died or out-migrated. Of the remaining 19,884 eligible women, 3,944 (19.8%) could not be contacted, 5,136 (25.8%) declined to participate in the annual population-based HIV testing in 2018, and 2,129 (10.7%) self-reported being on ART, thus resulting in 8,675 (31.9% of the resident population) who participated in the population-based HIV testing (Figure 1). Similar to men, randomization achieved balance in respect of HIV prevalence and sociodemographic variables in the arms with vs. without financial incentives, except for the area of residency (Table 1).

### HIV positive diagnosis by intervention arms among men and women

Among all 13,894 men ≥15 years resident in the 45 communities in 2018, the overall uptake of home-based HIV testing was 26.9% (1246/4624) in the financial incentive arms, and 17.5% (1621/9270) in the arms without the financial incentives. Of 20.6% men (n=2,867) who received HBHCT, 122 tested positive for HIV. In the ITT analysis, 1.4% (63/4624) in the financial incentive arms and 0.6% (59/9270) in the arms without the financial incentives tested HIV-positive using rapid HIV tests. The probability of a positive HIV diagnosis via HBHCT was about two times higher in the financial incentive arms among men, adjusting for the provision of the EPIC-HIV [adjusted risk ratio (aRR)=2.11, 95% CI: 1.36-3.28, p=0.001].

Among all 19,884 women ≥15 years living in the 45 communities, the overall testing coverage was 36.9% (2372/6433) in the financial incentive arms and 25.4% (3423/13451) in the arms without financial incentives. In the ITT analysis, compared to women in the arms without the financial incentives, the probability of uptake of HIV testing was 45% higher in the financial incentive arms (RR=1.45, 95% CI: 1.26-1.67, p<0.001). Of 5,795 (29.1%) who received HBHCT, 375 tested positive for HIV, including 60 (16.0%) women with previously unknown HIV status and 261 (69.6%) women newly diagnosed through the trial. Overall, 3.4% (217/6433) in the financial incentive arms tested HIV-positive, compared to 1.2% (158/13451) in the arms without financial incentives. The probability of a positive HIV diagnosis using home-based rapid HIV testing was 2.87 times higher among women in the financial incentive arms compared to the arms without the financial incentives (95% CI: 2.19-3.76, p<0.001).

### Effect of financial incentives on linkage to care at 6 weeks among men and women

Using individual clinical records ascertained from 17 clinics within the surveillance area, we confirmed that a total of 95 men initiated ART or resumed care within 6 weeks after the home visit. Among the 126 men who received positive (n=122) or discordant (n=4) HIV diagnosis via home-based HIV testing, 29 initiated ART or resumed care within 6 weeks (13 in the financial incentive arms and 16 in the non-financial incentive arms). Of 10,575 men who did not receive HBHCT, 1,166 were confirmed to be on ART, and 446 had interrupted care at the time of study visit. Among those who had interrupted care, 66 resumed care within 6 weeks (14 in the financial incentives and 52 in the non-financial incentive arms). In the ITT analysis, overall, 0.58% (27/4624) in the financial incentive arms and 0.73% (68/9270) in the arms without the financial incentives linked to care within 6 weeks after the home visit. The probability of linkage to care within 6 weeks did not differ significantly between the arms among men (aRR=0.78, 95% CI: 0.51-1.21).

On the other hand, a total of 198 women initiated ART or resumed care within 6 weeks after the home visit. Of the 395 women who received positive (n=375) or discordant (n=20) HIV diagnosis via home-based HIV testing, 77 initiated ART or resumed care within 6 weeks (45 in the financial incentive arms and 32 in the non-financial incentive arms). Among the 11,976 women who did not receive HBHCT, 2,743 were confirmed to be on ART, and 856 had interrupted care at the time of study visit. Among those with interrupted care, 121 resumed care within 6 weeks (38 in the financial incentives and 83 in the non-financial incentive arms). In the ITT analysis, 1.29% (83/6433) in the financial incentive arms and 0.85% (115/13451) in the arms without financial incentives linked to care within 6 weeks after the home visit. The probability of linkage to care within 6 weeks was significantly higher in the financial incentive arms among women (aRR=1.50, 95% CI:1.03–2.21, p=0.04).

The probability of linkage to care up to 1 year among men still did not differ in the financial incentive and non-financial incentive arms (Figure S2). As anticipated, among women, there was no additional gain in linkage to care up to 1 year in the financial incentive arms beyond the eligibility period of 6 weeks to receive the second financial incentive.

## DISCUSSION

In this study, we found that the small once-off financial incentive did not significantly improve linkage to care within the voucher eligibility period of 6 weeks in men whilst it substantially increased linkage to care by 47% among women. The provision of an immediate and tangible benefit via a financial incentive effectively reached those at higher risk, yielding a three-fold increase in HIV-positive diagnoses among both men and women. However, men encounter greater barriers in accessing care at clinics due to masculine norms, stigma, concerns about confidentiality, or the necessity to prioritize work over clinic visits, often resulting in delays in linking to care.^4116,19^ Our study finding suggests that the small once-off financial incentive is likely insufficient to overcome barriers that men face to link to care at clinics.

Several studies examined the efficacy of financial incentives on linkage to care and reported mixed results. In one study conducted in US metropolitan cities, financial incentives increased viral suppression and regular clinic attendance among men and women living with HIV but had no effect on linkage to care.^26^ In Mozambique, when financial incentives were provided in addition to comprehensive intervention strategy including health messages and appointment reminders via SMS messaging, there was no additional benefit of financial incentives in linkage to care.^25^

In our study, only 22% of men and 20% of women who tested HIV-positive via HBHCT across the study arms linked to care within 6 weeks of HBHCT, similar to the findings from other trials in South Africa. In the universal ART trial conducted in the north of the surveillance site (ANRS 12249 TasP cluster-randomized trial), only 36.1% of those tested HIV-positive via HBCHT linked to care within 3 months.^42^ Similarly, in the HPTN 071 (PopART) trial, only 31% of men and 34% of women diagnosed with HIV through HBHCT successfully linked to care within 3 months in South Africa.^17^ However, in Zimbabwe, when patients diagnosed with HIV at home were escorted to visit a clinic and link to care, over 85% successfully linked to care within just 30 days of HIV diagnosis.^43^ Similarly, in Botswana, facilitated and active linkage to care for HIV-positive individuals who were not receiving ART reduced the time to ART initiation by 81%, compared to the standard of care.^44^ Addressing the challenge of linkage to care, especially among men, would require a multitude of interventions simultaneously.

This study was conducted as a community-randomized clinical trial encompassing over 30,000 individuals nested within the ongoing population-based HIV surveillance. However, the overall population coverage of a once-off round of home-based HIV testing during the trial was relatively low at 21%, and as would be expected the HIV prevalence among those consenting to rapid-test for HIV was lower than the observed population prevalence in the communities. These led to a comparatively lower number of individuals who were eligible for linkage to care for ART initiation or re-engagement. Lastly, the intervention was delivered in a single round and might not be sustainable for routine implementation on a large scale. Policymakers might consider tailoring it for specific populations or settings in light of other competing interventions to optimize HIV response and treatment.

## CONCLUSION

In this cluster-randomized clinical trial, we found that during a single round of interventions to improve linkage to care, a small once-off financial incentive did not increase linkage to care among men during the eligibility period of 6 weeks. However, the provision of a small financial incentive significantly improved linkage to care among women during the same time period.

## Authors’ contributions

**Oluwafemi Adeagbo:** Conceptualization, Methodology, Software, Writing – Review & Editing. **Till Bärnighausen:** Conceptualization, Funding acquisition, Investigation, Methodology, Software, Supervision, Writing – Review & Editing. **Adrian Dobra:** Conceptualization, Formal analysis, Methodology, Writing – Review & Editing. **Dickman Gareta:** Data curation, Software, Writing – Review & Editing. **Maxime Inghels:** Conceptualization, Data curation, Formal analysis, Methodology, Validation, Writing – Review & Editing. **Hae-Young Kim:** Conceptualization, Data curation, Formal analysis, Methodology, Writing – Original Draft Preparation. **Thulile Mathenjwa:** Conceptualization, Investigation, Methodology, Project Administration, Software, Writing – Review & Editing. **Phillippa Matthews:** Conceptualization, Funding acquisition, Methodology, Software, Writing – Review & Editing. **Nuala McGrath:** Conceptualization, Funding acquisition, Methodology, Software, Writing – Review & Editing. **Jane Seeley:** Conceptualization, Funding acquisition, Methodology, Software, Writing – Review & Editing. **Maryam Shahmanesh:** Conceptualization, Funding acquisition, Methodology, Software, Writing – Review & Editing. **Frank Tanser:** Conceptualization, Funding acquisition, Investigation, Methodology, Software, Supervision, Writing – Review & Editing. **H Manisha Yapa:** Conceptualization, Methodology, Writing – Review & Editing. **Thembelihle Zuma:** Conceptualization, Methodology, Writing – Review & Editing.

## Data Availability

The data underlying the results presented in the study are available from the AHRI Data Repository (https://data.ahri.org) for researchers who meet the criteria for access to confidential data and sign on the agreement according to the AHRI's protocol for data sharing. The authors confirm they did not have any special access or privileges that others would not have.

https://data.ahri.org/index.php/home

## Acknowledgement

We thank community members for their continued support and participation in the Africa Health Research Institute’s Health and Demographic Surveillance System (HDSS), and the AHRI population, clinical, laboratory and data management teams – especially, Kobus Herbst, Thobeka Mngomezulu, Siyabonga Nxumalo, Keabetswe Dikgale, Jaco Dreyer, Theresa Smit, Sphephelo Dlamini and his team, Sithembiso Luthuli, and Patrick Gabela.

## Appendix

**Figure S1.**
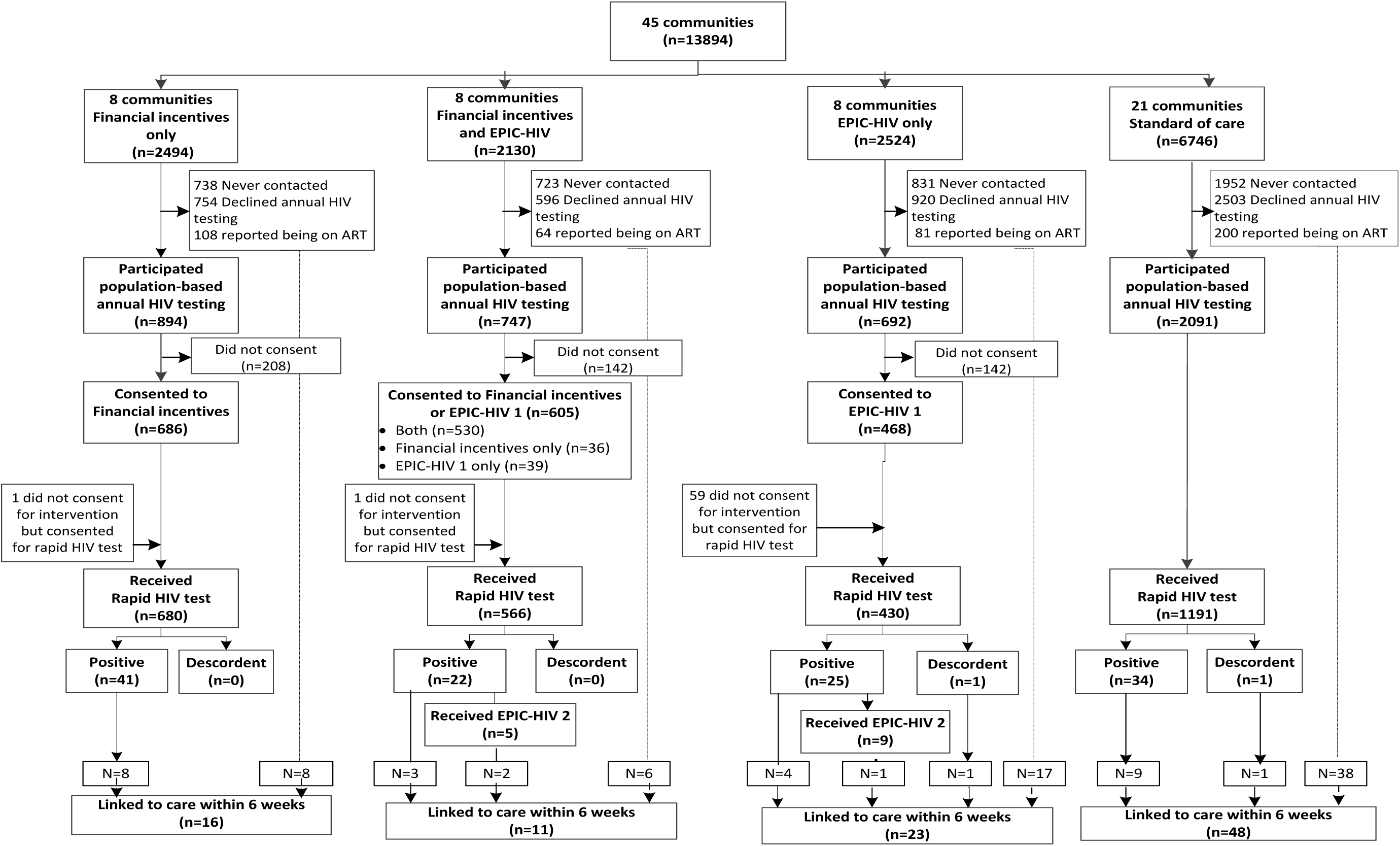
Flow diagram for the HITS cluster-randomized controlled trial and linkage to care within the voucher eligibility period of 6 weeks of a home visit among men. Flow diagram shows individual flow through each stage of the HITS trial by intervention arms. The dashed line indicates linkage to care within 6 weeks among those who were never contacted, declined annual HIV testing, or did not consent for interventions. Abbreviation: EPIC, Empowering People through Informed Choices for HIV.

**Figure S2.**
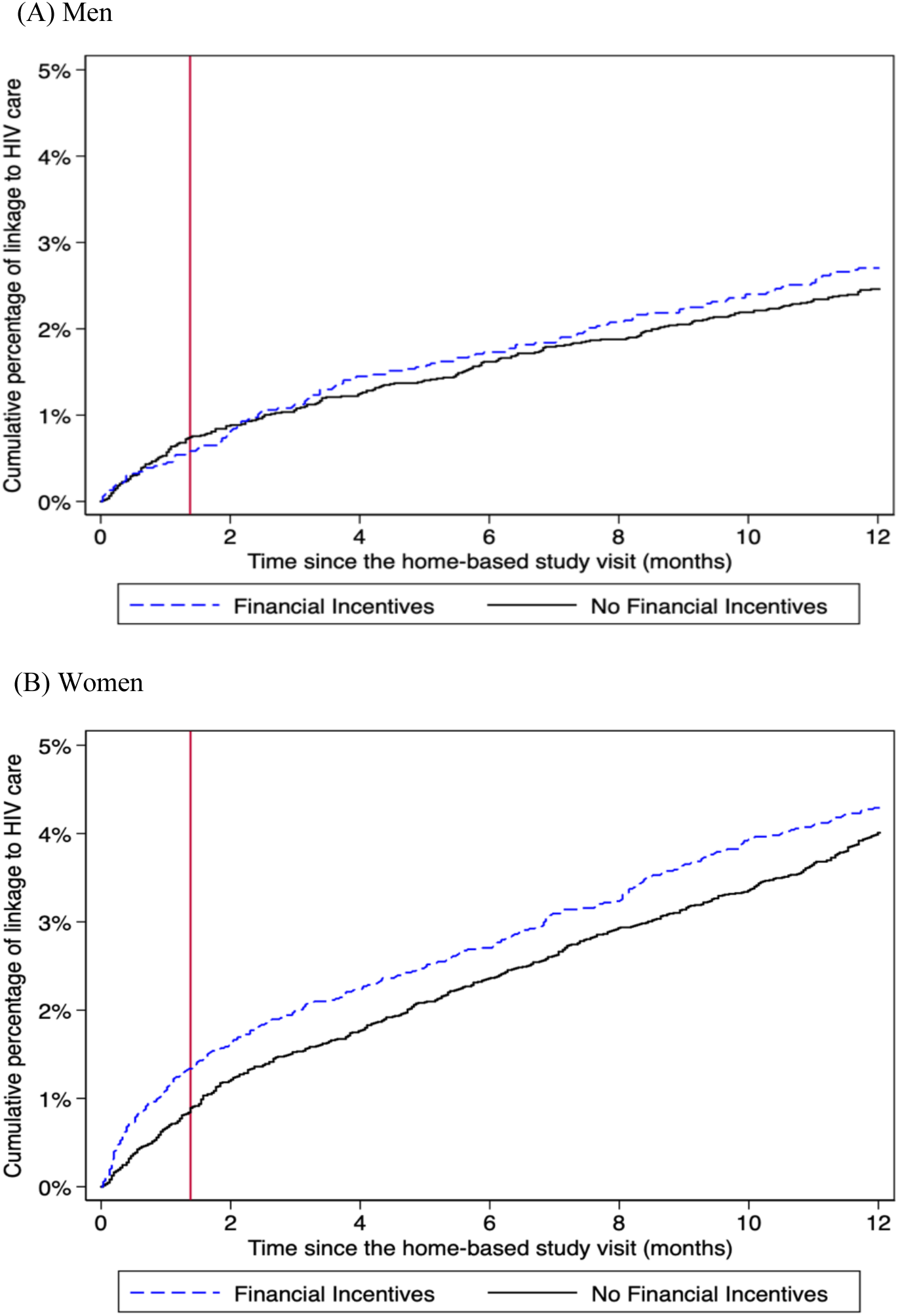
Kaplan Meir curves for linkage to HIV care by 1 year after the home-based study visit among (A) men and (B) women. The solid black lines represent the arms without financial incentives, and the dashed blue lines the financial incentive arms. The red line indicates the voucher eligibility period of 6 weeks to receive the second financial incentive when linked to care.

